# Factors associated with teenage pregnancies during the Covid-19 period in Pakwach district, Northern Uganda: a case-control study

**DOI:** 10.1101/2023.09.13.23295521

**Authors:** Jimmy Patrick Alunyo, David Mukunya, Agnes Napyo, Joseph KB Matovu, David Okia, Benon Wanume, Francis Okello, Ally Hassan Tuwa, Daniel Wenani, Ambrose Okibure, Godfrey Omara, Peter Olupot-Olupot

## Abstract

**Background:** Teenage pregnancy rates have globally decreased over the years, but remain high, especially in low- and middle-income countries (LMICs). Among girls aged 15-19, teenage pregnancy remains the leading cause of death and a significant barrier to education and productivity. Its prevalence underscores concern about the sexual and reproductive health of youth. However, limited data exist regarding factors contributing to its rise during the COVID-19 pandemic in Uganda. This study explores the factors associated with teenage pregnancy in Pakwach district during this period.

**Methods:** We conducted a matched case-control study, enrolling 362 teenage girls aged 10-19 years, divided into two groups: 181 pregnant teenagers and 181 not pregnant teenagers. We collected exposure data from both groups using questionnaires to evaluate factors associated with teenage pregnancy. The study period covered March 2020 to January 2021, coinciding with lockdown measures.

**Results:** During the COVID-19 lockdown, teenage pregnancies were only associated with having exclusively female peers (AOR 3.0, 95% CI: 0.1-104.4). Conversely, having a Radio/TV at home (AOR 0.2, 95% CI: 0.1-0.6), age at first sexual encounter (AOR 0.1, 95% CI: 0.03-0.9), considering teenage pregnancy as sexual abuse (AOR 0.1, 95% CI: 0.02-0.4), feeling comfortable asking questions during consultations (AOR 0.5, 95% CI: 0.2-1.3), and ensuring sufficient privacy during consultations were protective against teenage pregnancy.

**Conclusion:** The factors contributing to increased teenage pregnancies during the COVID-19 pandemic were consistent with long-standing contextual factors associated with teenage pregnancy. The lockdown environment may have slightly exacerbated these factors, but no direct association was observed. Only having female peers was linked to teenage pregnancy during the lockdown. Conversely, having access to a radio/TV at home and other healthcare system-related factors offered protection. Therefore, interventions should prioritize providing comprehensive information on the risks of teenage pregnancy during any lockdown scenario.

## Background

Teenage pregnancy is a global public health problem, with a preponderance in LMICs where it accounts for 95% of all cases, and Sub-Saharan Africa included. The pooled prevalence of teenage pregnancy in Sub-Saharan African countries stands at 18.8%(4), with significant variation across regions. The East African region has the highest prevalence of 21.5%, followed by 20.4% in southern Africa, 17.7% in west Africa, 15.8% and 9.2% in Central Africa, and northern Africa respectively (5). The available data shows significant variation between the countries of Africa. For instance, in west Africa, Liberia has the highest prevalence at 38.9% NDHS 2013, followed by Gabon at 38.0% NDHS 2012 Mali in 36.3% NDHS 2018. In east Africa, Uganda has the highest prevalence of 26.1% UDHS 2016, followed by Tanzania (25.1%) NDHS 2015-2016, then Kenya (18%) NDHS 2014, Burundi (7.9%) NDHS 2016-2017, and Rwanda the least (7.2%) NDHS 2015. And in southern Africa, Angola has the highest prevalence of 39.4% NDHS 2015-2016, followed by Namibia (21%) NDHS 2013, Lesotho (19.8%) NDHS 2014 and south Africa at 16.9% NDHS 2016, according to a systematic review on prevalence of first teenage pregnancies across African countries (6).

The factors responsible for teenage pregnancies in the African continent has previously be been investigated and factors likes being a victim of sexual abuse, risky sexual behaviors such as early sexual initiation, and non-use of contraceptives among others were found as key predicted of teenage pregnancies (6) (10).

Teenage pregnancies carry significant public health importance due to their complications, which are the leading cause of mortality among girls aged 15-19 globally. Physiological and socio-economic factors contribute to various complications associated with teenage pregnancy. These complications include preterm birth, low birth weight, anemia, malnutrition, high blood pressure, and emotional and social challenges commonly observed in pregnant teenagers. These complications are influenced by factors such as lack of parental care, inadequate nutrition, underdeveloped reproductive systems, and economic difficulties, particularly intensified by the COVID-19 pandemic.

The impact of teenage pregnancy is substantial, affecting young mothers worldwide and presenting a range of challenges such as lower educational attainment, reproductive health issues, higher fertility rates, decreased economic earnings, domestic violence, and limited opportunities. Furthermore, it has long-term effects on infants, increasing the risk of mental disabilities and neurological problems.

With the emergence of the novel coronavirus, the impact of the COVID-19 pandemic on the adolescent population in terms of teenage pregnancy and associated risk factors remains uncertain. In Uganda, before the outbreak of the pandemic, the trends was decreasing at a stagnated rate, despite its high prevalence at 25% among teenagers in the year 2016(15). This prevalence for the east African region is the highest by far. We hypothesized that the prolonged lockdown in response to the pandemic escalated the risk of teenage pregnancies in Uganda. Therefore, we aimed to provide COVID-19 context-specific factors associated with teenage pregnancies during COVID-19 in Pakwach district Uganda.

## Methods and Materials

### Study setting

The study area was Pakwach District, located in the West Nile Region, northern Uganda. According to the Uganda Bureau of Statistics figures, the district had a total population of 158,037 in 2014 and was projected to increase to 181,400 by 2018, and 51.7% of this are females. Pakwach has six sub-counties (Pakwach Town Council, Panyango, Alwi, Pakwach Sub County, Wadelai and Panyimur), with 30 wards and 414 villages. From these six sub counties, three were selected randomly to be included in this study (PTC, Panyanago and PSC). We visited all the health facilities in the selected sub counties and all records of first ANC services utilization for girls aged 10-19 years were accessed from the registers. A sampling frame was developed from this list and proportionate allocative technique was used to select participants. Simple random sampling was used to selected cases for inclusion.

### Study design

The study design was a case control study. We accessed the ANC registers available at the health facilities of the selected sub-counties. Records of pregnant teenagers aged 10-19 were extracted. This became the sampling frame, where cases were selected randomly. The research assistants visited their homes and interviewed them.

The control group consisted of age-matched teenagers who were nominated by the cases. These controls confirmed that they had never been pregnant and had no known history of pregnancy. Matching in this study was performed based on factors such as educational level, school attendance at the time of conception, neighborhood (for girls not in school during data collection), age, and location. The purpose of matching was to control for confounding. Each case was asked to provide the names of five peers of the same age who attended their school (not necessarily from the same class), or who lived in the same neighborhood if they were not in school. The last two names provided as potential controls were approached first and invited to participate in the study. Importantly, none of the subjects declined to participate in the study.

### Target population

The target population were teenage girls aged 10-19 years. They were either pregnant or not pregnant teenagers who had attended ANC or access health services in the three selected sub-counties, from March 20th, 2020, to January 2021.

### Sample size and Sampling procedures

The sample size for this study was calculated using the Epi tools calculator used for Sample size estimation for Case-control studies. Three hundred sixty-two (362) participants were included in this study using the specification explained in the paragraph below. The expected proportion of teenage pregnancy in the control group, as reported in the (UDHS 2016), was 0.224. We used an assumed Odds ratio of 2, aiming for a power of 0.85 and a confidence interval of 0.95. A total of 362 participants were included in the sample, divided equally between cases (181) and controls (181), as shown in the table below. The primary sampling frame for the study consisted of the OPD register, from which records of teenage girls aged 10-19 accessing antenatal care services were obtained. A sampling frame was then created, and a systematic random sampling method was employed to select the cases. The cases were tracked and interviewed within the communities. At the community level, each case was asked to provide the names of five peers of the same age who attended their school (not necessarily from the same class) if they were attending school, or who resided in the same neighborhood if they were not in school. These identified peers were then selected for interviews as a control group.

### Data collection

A structured questionnaire designed in electronic format (Kobo Toolbox) was used to interview participants. The development of this tool was informed by WHO Illustrative questionnaire developed by John Cleland (Cleland, 2001) for interview surveys with young people. This standard tool was adjusted to fit the local context. It had questions on socio demographic characteristics, economic factors, teenage factors, and health system factors, environmental factors, and cultural factors.

### Data Management

The data was downloaded in Excel and subsequently checked. It was then exported to Stata 15 for cleaning, coding, and analysis. Initially, at the univariable level, frequencies and percentages were calculated and presented in tables and charts. In the bivariable analysis, each risk factor was tested against the dependent variable, and the results were displayed in tables as odds ratios and Pearson chi-square p-values for both cases and controls.

To determine the strength of association between the exposure variables and the main outcomes (Pregnant/Not Pregnant), logistic regression was utilized. In this analysis, conventional logistic regression was employed since matching was carried out to control confounding variables but was not the primary focus of the analysis. The primary objective of this analysis was to estimate the association between the exposure and outcome.

The results were reported as adjusted odds ratios along with corresponding 95% confidence intervals (CIs)

### Ethics approval and consent to participate

Ethical consideration was sought from Busitema University Higher Degree committee and Mbale Regional Referral REC. The approval for this study, reference number (MRRH-2021-75: A) was Granted by MRRH-REC accredited by UNCST, registration number UG-REC-001 through expedited review on 13/08/2021. Informed consents were sought from the study participants before starting data collection. Additional written informed consents were also obtained from the parent/ guardian in cases where we sampled a child and further authoritative clearances to conduct the study were also sought from the District Health Officer, Chief Administrative Officer, District Education Officer, Resident District Commissioner, Facility In charges and the LC1 chairpersons in all Villages. All the stipulated COIVID-19 standard operating procedures (SOPs) set by the government were adhered at all levels to protect the study subjects from any harm of contracting the COVID-19 virus during data collection.

## Results

### Demographic profile of participants

A total of 362 adolescent girls, with a mean age of 17.7 years (Std. Err 0.063, 95% CI = 17.575, 17.823), participated in the study. Approximately 49.2% of the participants were recruited from Pakwach sub-county, and the majority of them (99.7%) fell into the older teenage category (15-19 years). Across all groups, the literacy level was high, with 95.8% of the girls being able to read and write. Additionally, the majority (82.6%) were attending primary school. A significant proportion (91.7%) of the participants were unmarried.

**Fig 1.**
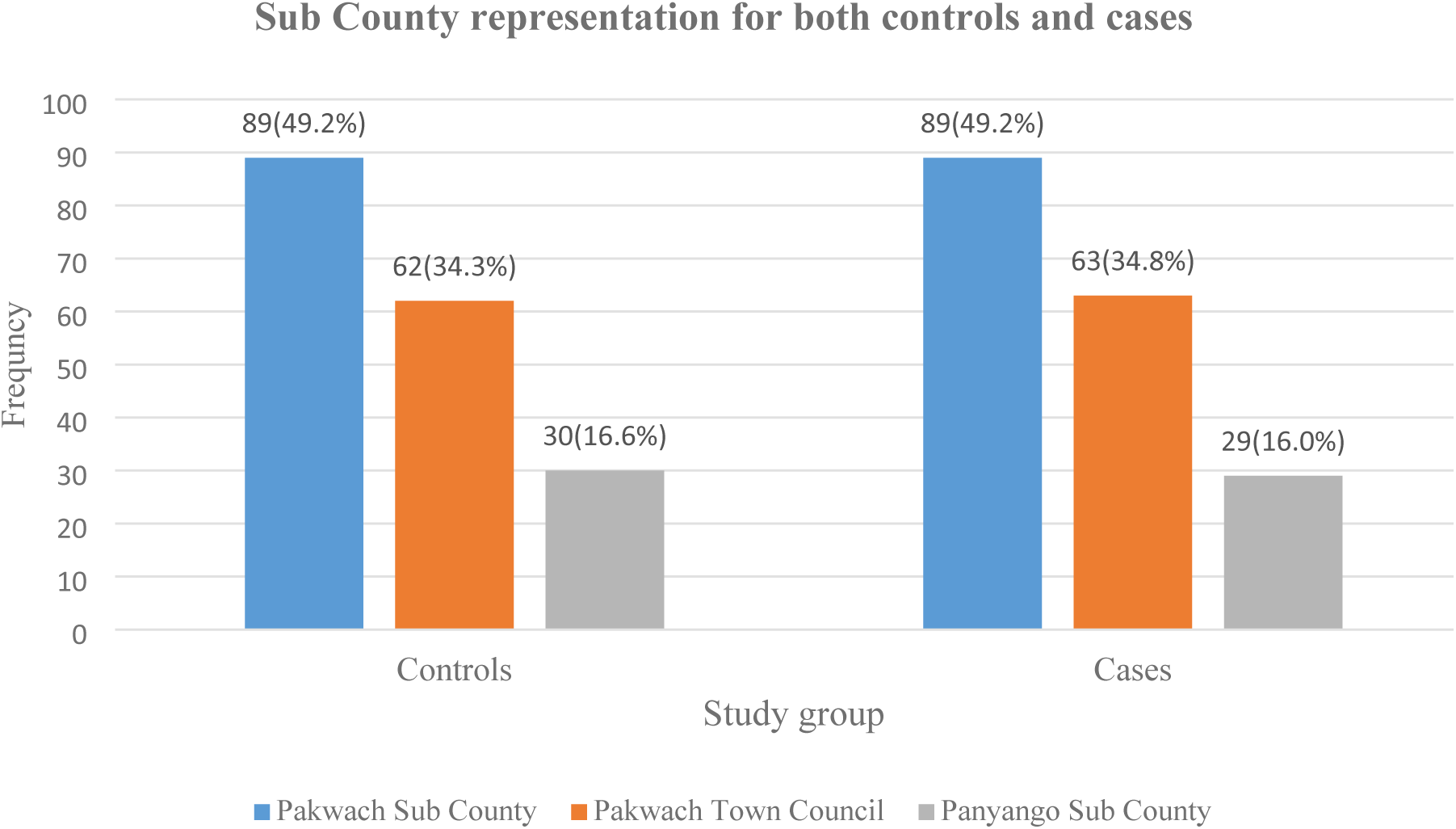
Showing a graphical presentation of Sub counties

**Fig 2.**
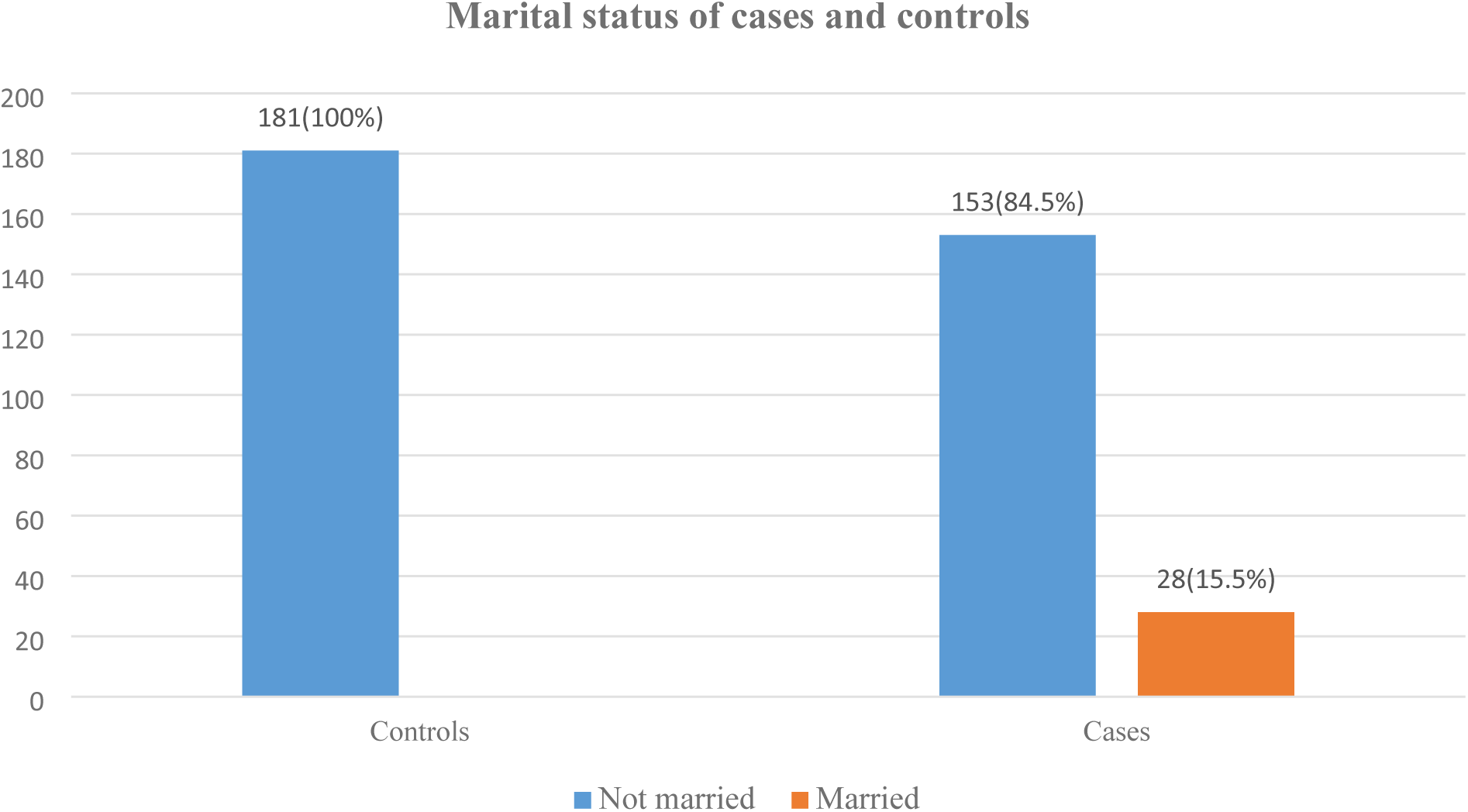
Shows the marital status of our participants.

**Fig 3.**
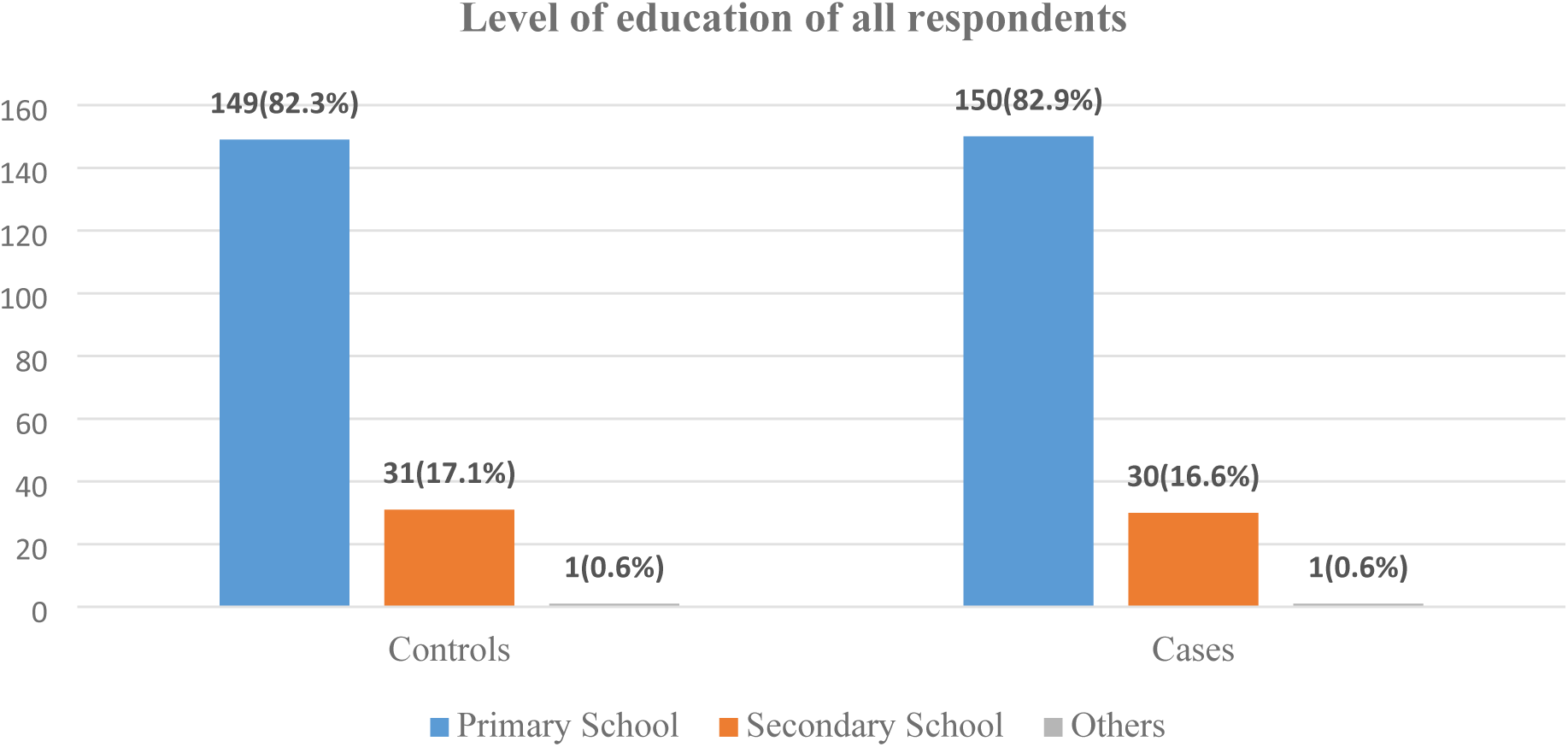
Shows the level of education of all participants.

**Table 2**. The results for age categorization, literacy level, health facility visits, and the type of services requested by teenagers during the COVID-19 lockdown are presented. Throughout the lockdown period, 68.2% of the participants visited a health facility. Most of these visits (99%) were recorded in government health facilities. A significant portion of the teenagers (75%) made between 2 to 4 visits to the health facilities. In terms of service demand, there was a high demand for pregnancy services (97.6%).

**Table 1.**
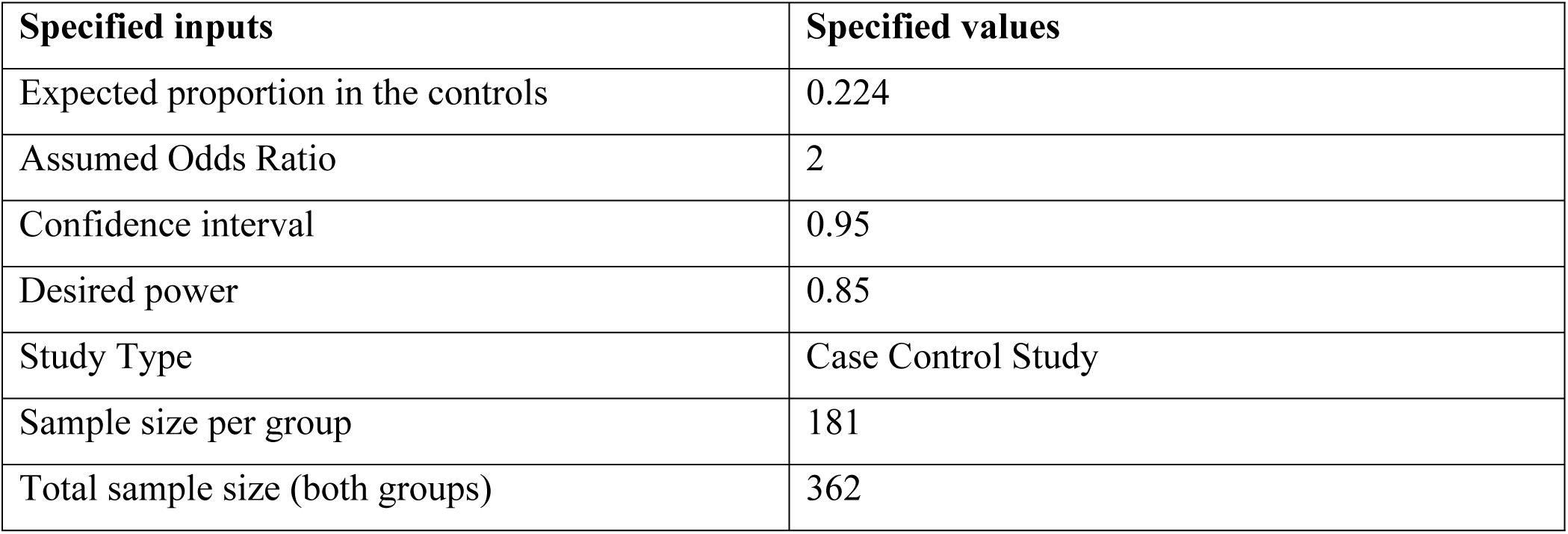
Shows the breakdown of the sample size calculation for this study.

| **Specified inputs** | **Specified values** |
| --- | --- |
| Expected proportion in the controls | 0.224 |
| Assumed Odds Ratio | 2 |
| Confidence interval | 0.95 |
| Desired power | 0.85 |
| Study Type | Case Control Study |
| Sample size per group | 181 |
| Total sample size (both groups) | 362 |

**Table 2.** Showing Age Categorizations, Literacy level, Health facilities visits, and type of services requested by Teenagers.

| Variable | Frequency n=362 | Percentage (%) |
| --- | --- | --- |
| Age |  |  |
| Young Teenage (10-14 yrs) | 1 | 0.3 |
| Older Teenage (15-19 yrs) | 361 | 99.7 |
| Literacy level | 346 | 95.8 |
| Health facility visit during Lockdown | 247 | 68.20 |
| Number of Visit |  |  |
| <2 times | 28 | 11.30 |
| 2-4 times | 188 | 76.10 |
| >=5 times | 31 | 12.60 |
| Types of services Requested at the facility |  |  |
| Contraceptives | 113 | 45.7 |
| Pregnancy services | 121 | 49% |
| Facility Type |  |  |
| Government | 244 | 98.80 |
| Private | 3 | 1.20 |
| Health information | 230 | 93.10 |
| The attending clinician talked about the following services |  |  |
| Condom use | 160 | 64.8 |
| Contraceptives | 161 | 65.2 |
| Pregnancy | 241 | 97.6 |
| Attending clinician receptive | 235 | 95.10 |
| Received the services/care one went looking for | 232 | 93.90 |
| Felt comfortable enough to ask questions | 136 | 55.10 |
| Consultation Questions answered adequately | 169 | 68.40 |
| There was enough privacy | 170 | 68.80 |

Almost all teenage girls (99%) who visited health facilities during the COVID-19 lockdown reported receiving health information. The most prevalent discussions focused on pregnancy prevention, with a particular emphasis on contraceptives and condom use. Generally, the attending clinicians provided a good reception for our participants. More than half of the girls (68.8%) reported having privacy during their consultations, and 55.1% felt comfortable asking questions. Overall, 68.4% stated that their questions were adequately answered by the attending clinicians. Importantly, nearly all the participants (93.9%) confirmed that they received the services they were seeking at the health facility.

**Table 3**. Presents location and social demographic factors for teenage pregnancies. In Table 3, we see that most pregnancies (99%) occurred in older teenagers aged 15-19 years, and about 92% of them were still living at home with their parents and not married. There was high literacy level (92%) among our participants, and this was statistically significant at P< 0.001.

**Table 3.**
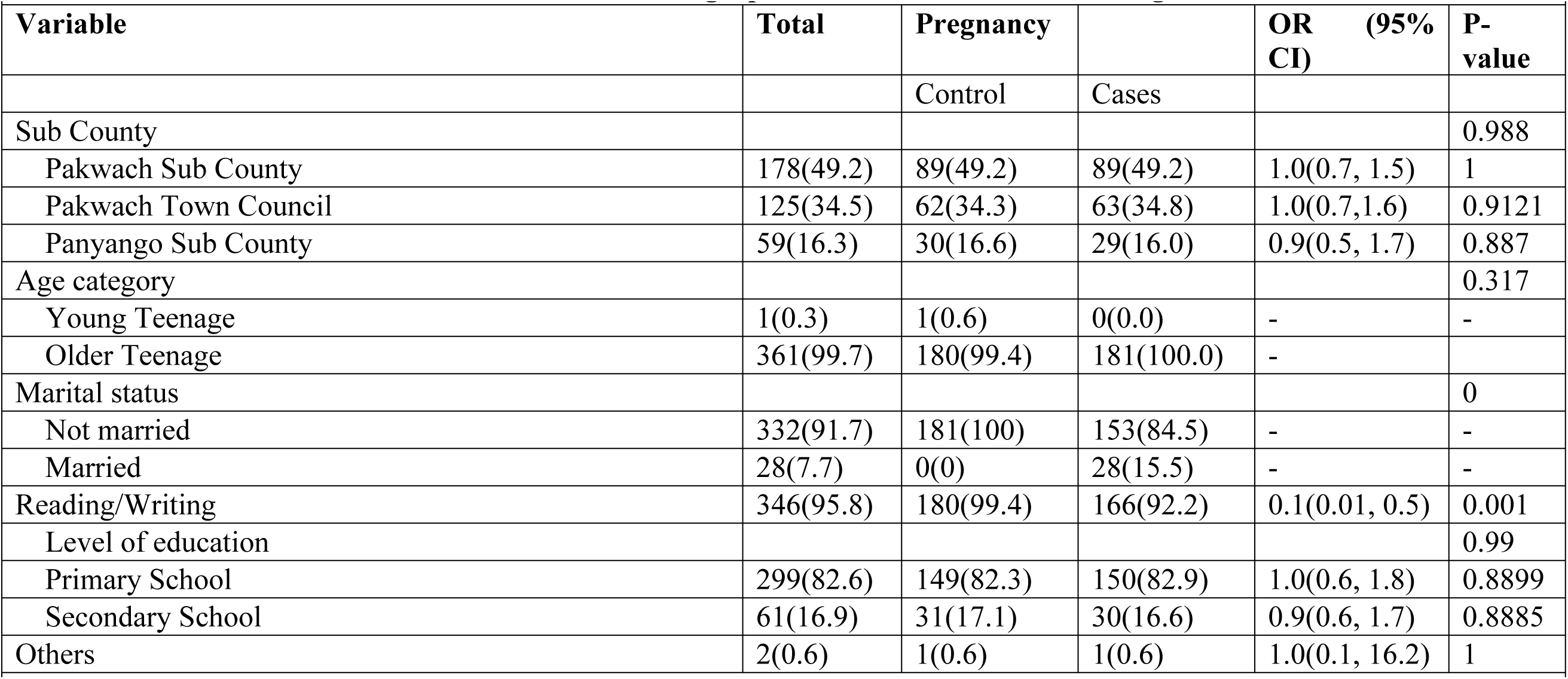
Bivariate results for location and social-demographic characteristics of the teenagers.

**Table 4**. Presents results on health facility visits and the type of services requested by teenagers during the COVID-19 lockdown. Health facility visits were more among the pregnant group (79%). These visits strongly predicted the likelihood of getting pregnant, and this was associated with 2.9 times higher odds of pregnancy among those who visited the health facility. This was statistically significant at P<00001. However, the frequencies of those visits were significantly associated with reduced likelihood of getting pregnant at P<0.0077.

**Table 4.**
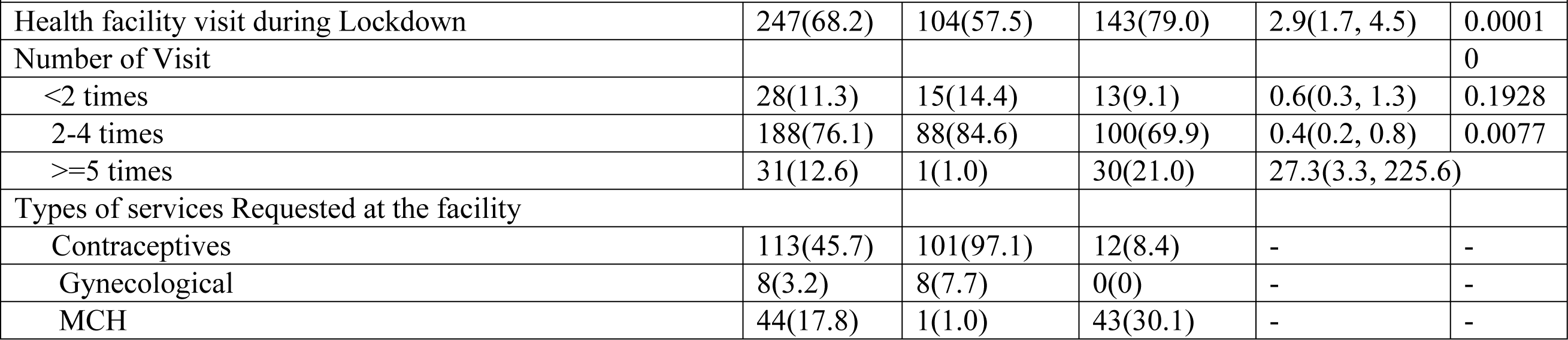
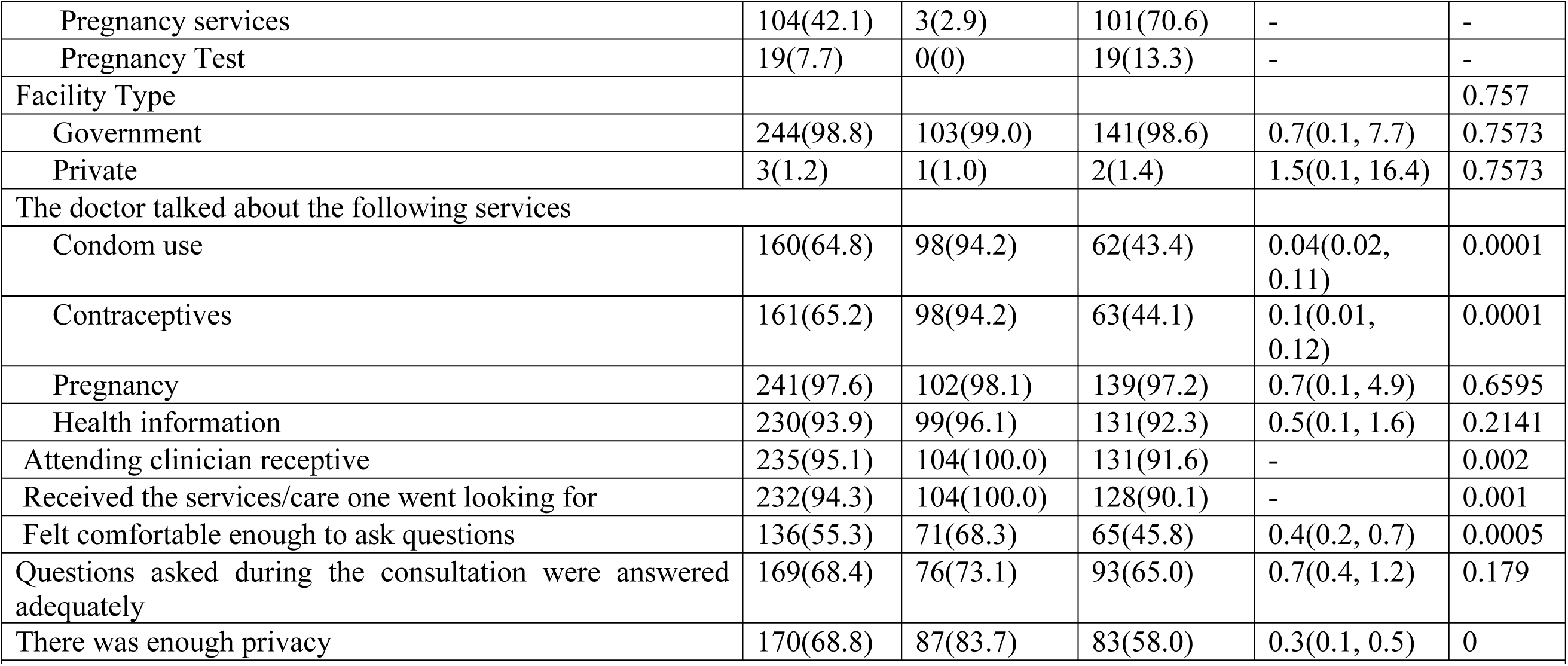
Bivariate results for health facility visits and type of services requested for by teenagers.

Attending more health talks on contraceptive use (64.8%) and condom use (65.2%) was also associated with less likelihood of getting pregnant P<0.0001. More interestingly, getting good reception at the facility from the attending clinician, also had a significant association with not getting pregnant (P<0.002). But overall, 90% of the teenagers who visited the health facility received the services they went looking for at the facility, and this was also significant at P<0.001. Other health system factors like feeling comfortable to ask questions during consultation, questions being answered adequately and having enough privacy during consultation also had significant relationship with not getting pregnant.

**Table 5**. Presents the type of peers, type of friends, and pregnancy prevention methods used by teenagers. Almost all our participants (96.4%) had friends and female sex were the most preferred type of peers by our teenagers (86.2 %). Being friends with a mature individual and having only girls as friends, were associated with getting pregnant. This was statistically significant at P<0.0001. Having older siblings at home was however, associated with 0.5 times less likelihood of getting pregnant, but being in a relationship was associated with 5.8 times high likelihood of getting pregnant. Most teenagers reported having their first sexual intercourse between the age 15-19 years, and over 60% of them regretted it. This had a significant association with teenage pregnancy at P<0.0024. More than half of our participants (59.4%) were also using pregnancy prevention methods, with condom use being the most preferred method. This was associated with a high likelihood of getting pregnant [COR 1.6, 95% CI= (1.1, 2.5)]. Sexual abuse (9.4%) and Sex trade (10.5%) also strongly predicted pregnancy P<0.000.

**Table 5.**
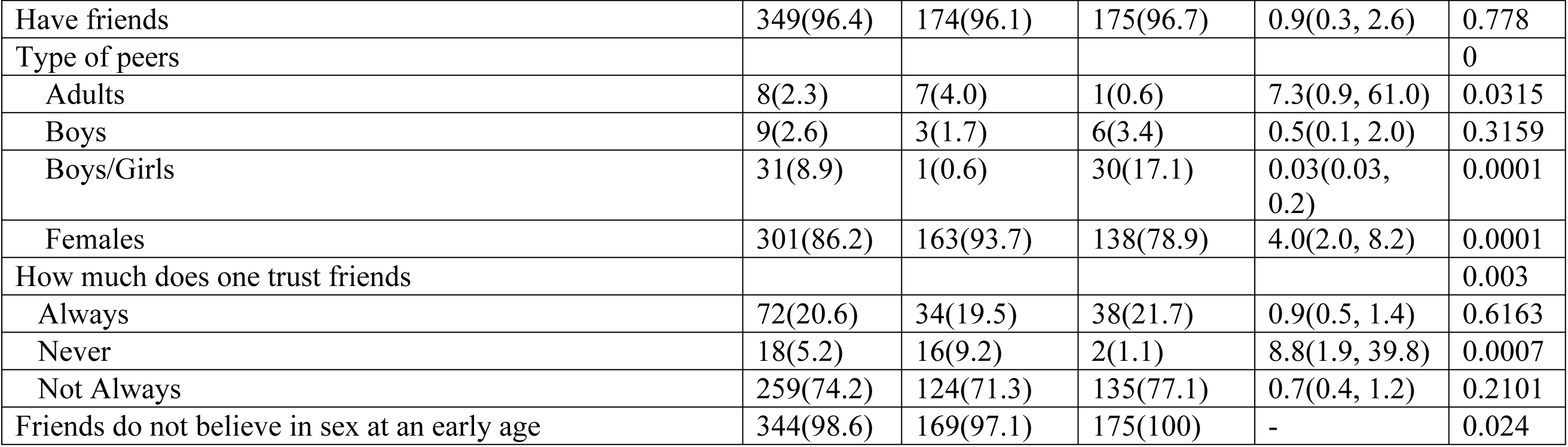
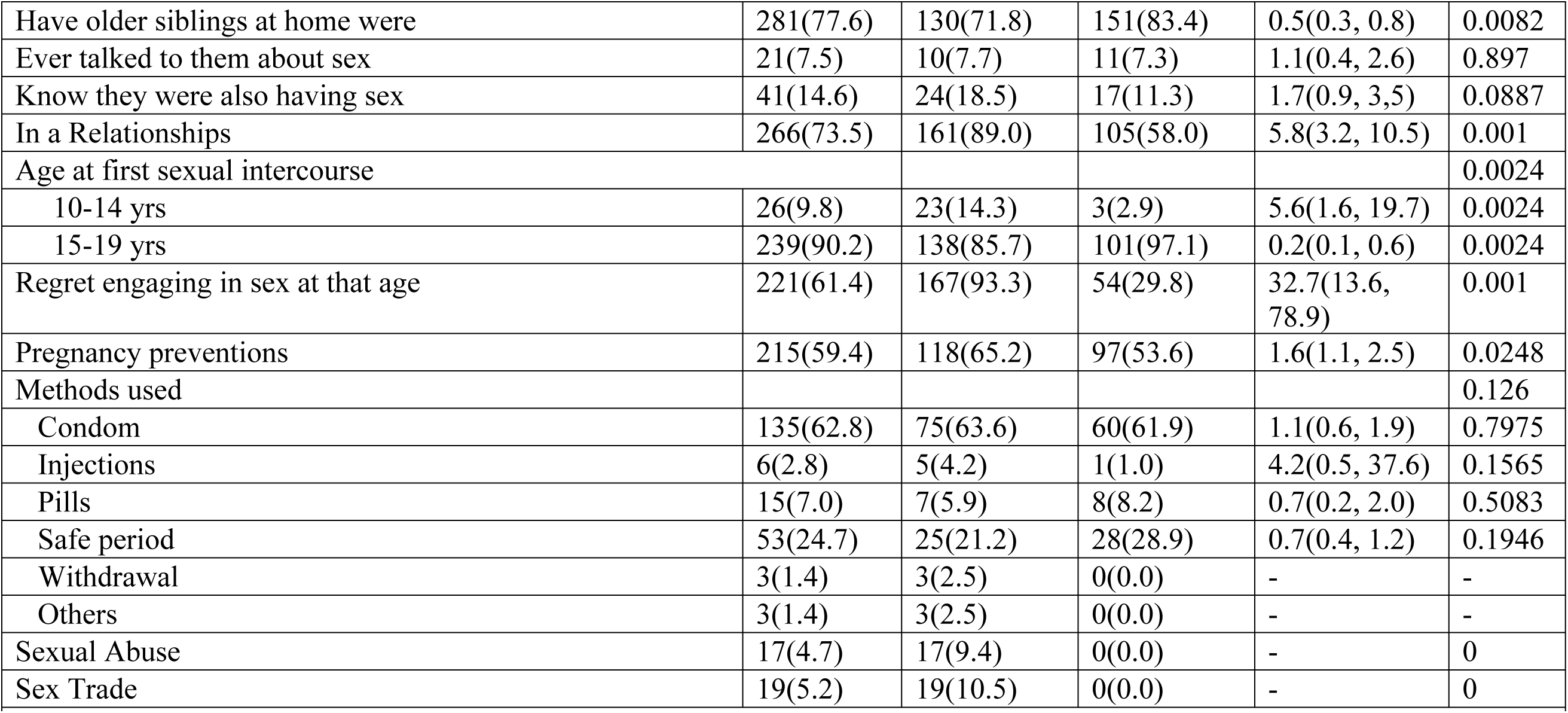
Bivariate results for the type of peers, type of friends, and pregnancy prevention methods used by teenagers.

**Table 6**. Presents socioeconomic characterization and teenagers’ perception of pregnancy. Ownership of a radio and television during the lockdown was protective against teenage pregnancy P<0.0001. The time of returning home from places of watching TV/listening to the radio also predicted teenage pregnancy, with Afternoon time representing a high likelihood of pregnancy [OR 4.7, 95% CI= (0.9, 23.6)].

**Table 6.**
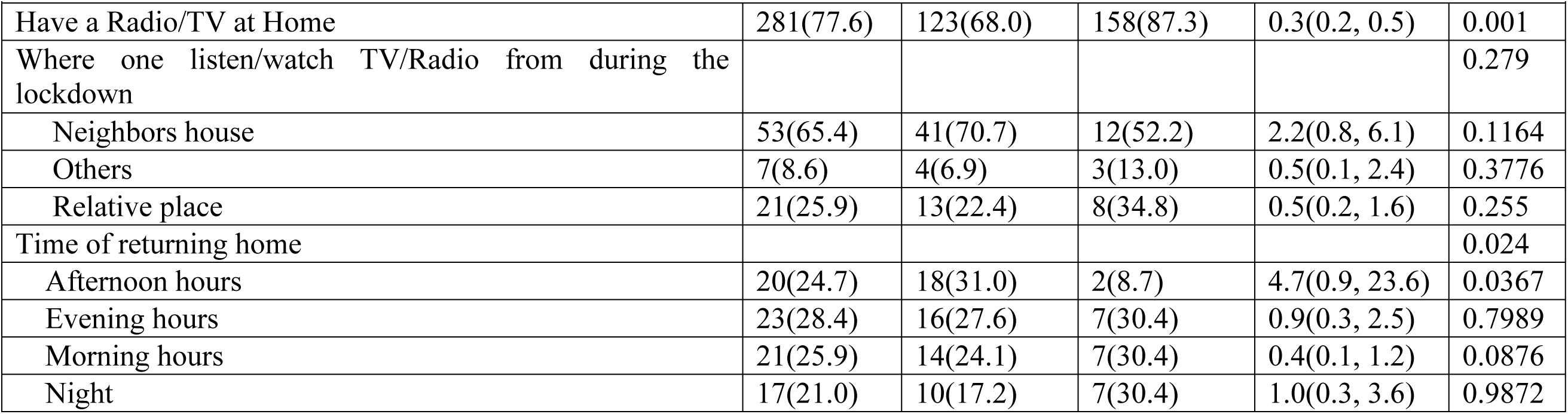
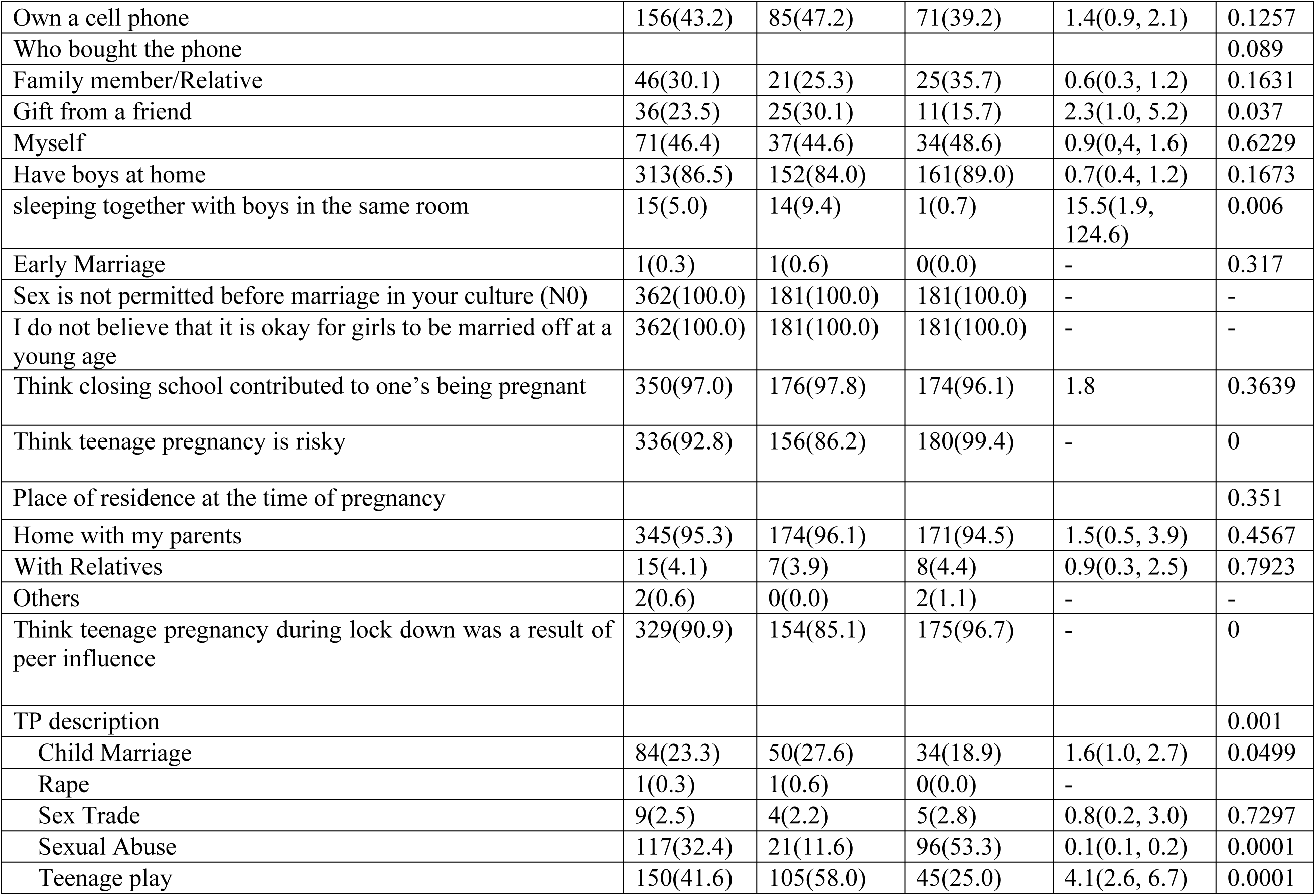
Socioeconomic characterization and teenagers’ perception of pregnancy.

Acquiring phones as gifts from friends was also associated with a 2.3 times higher likelihood of getting pregnant. Thinking that teenage pregnancy was risky was also associated with less likelihood of getting pregnant at P<0.0001. Residing at home with parents also represented 1.5 times higher odds of getting pregnant.

### Outcomes of teenage pregnancies during the COVID-19 lockdown in Pakwach district

**Table 7** Shows that the majority 170/181 (94.4%) of the pregnancies were unintended, and 95% were living outside the marital union. Health facility delivery was high at 95 %. Of the 181 girls who got pregnant, 88.3% (N=159) had a live birth, 11.1% (N=20) were neonatal death, and only 0.6 % (one) was an abortion (Figure 4). There was also a high school dropout (71.3%) among our participants.

**Table 8.** General characterization of pregnancies among teenagers.

| Variables | Frequency<br>N=181 | Percentage (%) |
| --- | --- | --- |
| **Pregnancy Cases** | 181 | 50.0 |
| **Planned/Unplanned** |  |  |
| Planned | 10 | 5.6 |
| Unplanned | 170 | 94.4 |
| **Married to the father of the child** | 33 | 18.2 |
| **School return after delivery** |  |  |
| Dropped out | 129 | 71.3 |
| In school | 52 | 28.7 |

**Figure 4.**
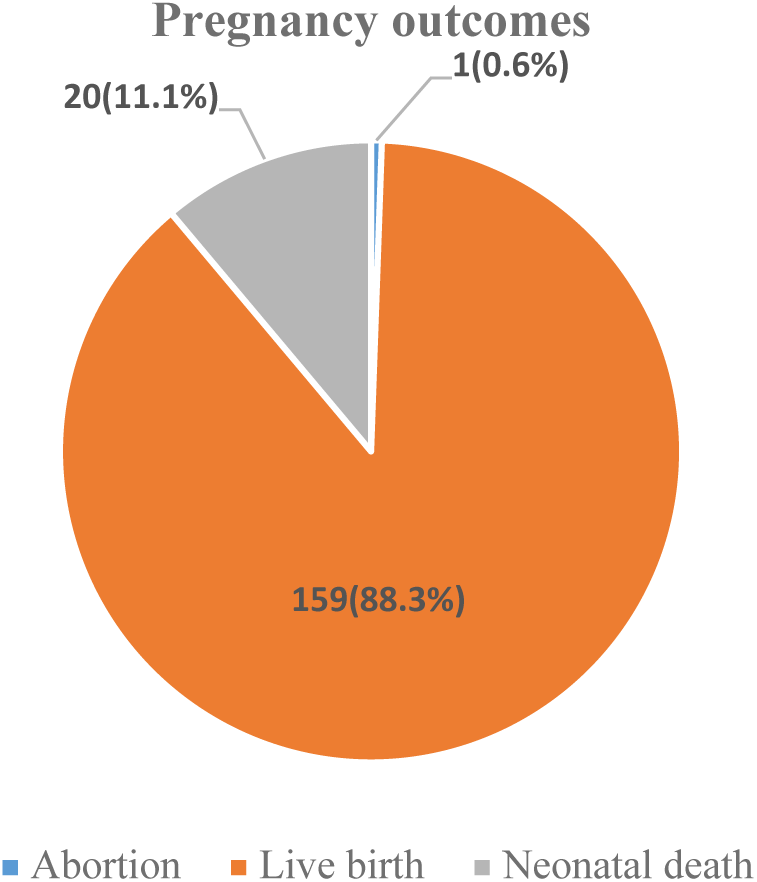
A pie chart showing pregnancy outcomes among teenagers.

**Figure 5.**
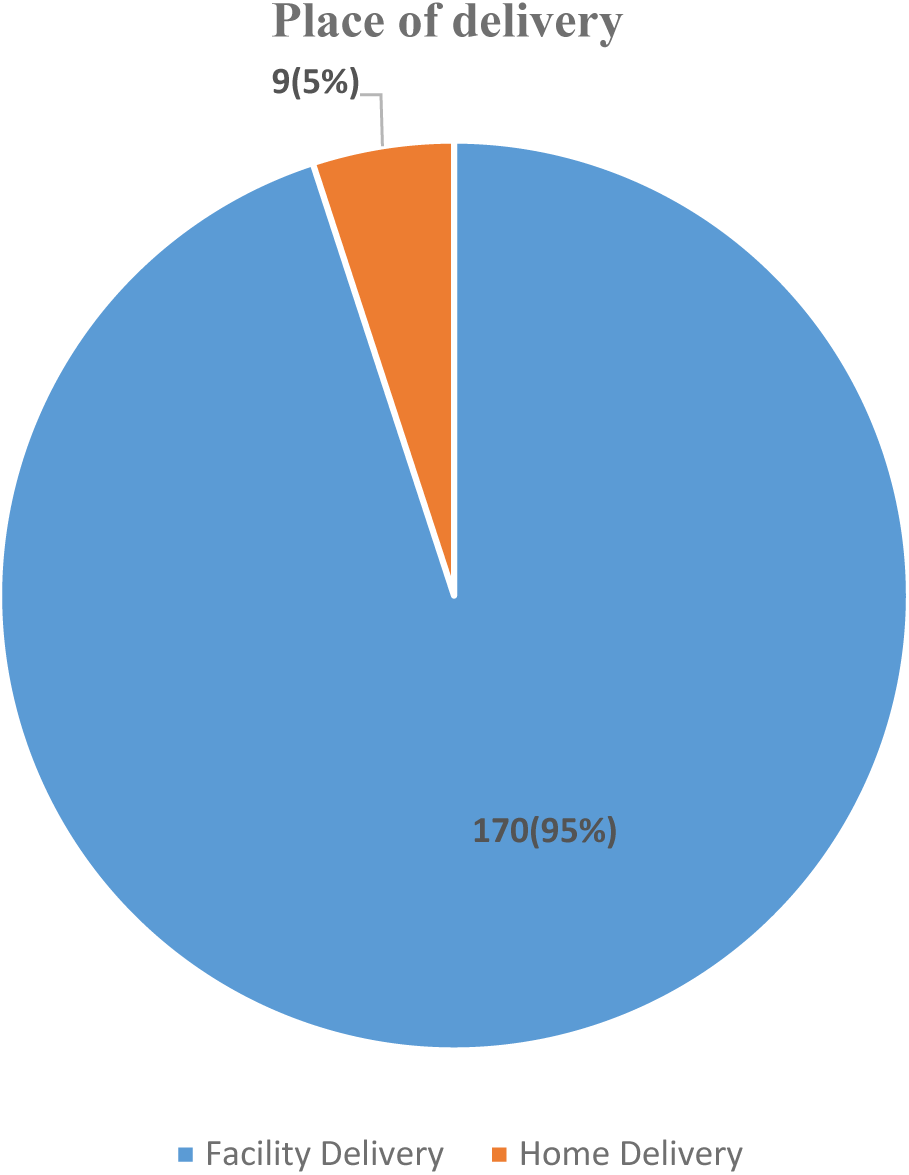
A Pie chart showing the place of delivery chosen by teenagers.

**Table 9** Presents the result of pregnancy birth outcomes versus Place of delivery. Nearly one fourth (22.2%) of neonatal death happened at home and only 10.5% at the health facility. Health facility delivery was associated with more (96.2%) live birth than home delivery, characterized by high neonatal death abortion. Delivery place strongly predicted neonatal death and abortion at P<0.0001.

**Table 9.** Pregnancy birth Outcomes Vs Delivery place.

| Pregnancy outcome | Place of delivery |  |  |  |
| --- | --- | --- | --- | --- |
|  | Facility Delivery | Home Delivery | Total | P-value |
| **Abortion** | 0(0.0) | 1(100.0) | 1 | 0.0001 |
| **Live birth** | 153(96.2) | 6(3.8) | 159 |  |
| **Neonatal death** | 18(90) | 2(10.0) | 20 |  |
| **Total** | 171(95.0) | 9(5.0) | 180 |  |

### Multivariate results

After adjusting for all other factors as shown in Table 7, pregnancies in teenagers during the COVID-19 lockdown in Pakwach district was only found to be associated with having only females as friends by teenagers [AOR 3.0, (0.1, 104.4)]. However, viewing teenage pregnancy as sexual abuse [AOR 0.1, 95% CI = (0.02, 0.4)] having a Radio/TV at home [AOR 0.2, 95% CI=(0.1, 0.6)], age at first sexual encounter (15-19 years) [AOR 0.1,95% CI=(0.03, 0.9)], feeling comfortable to ask questions during consultation [AOR 0.5,95% CI=(0.2, 1.3)], having enough privacy during consultation [AOR 0.3, CI= (0.1, 0.9)], and using pregnancy preventions [AOR 0.04, CI= (0.01, 0.2)] were found less associated with the likelihoods of getting pregnant.

**Table 10.**
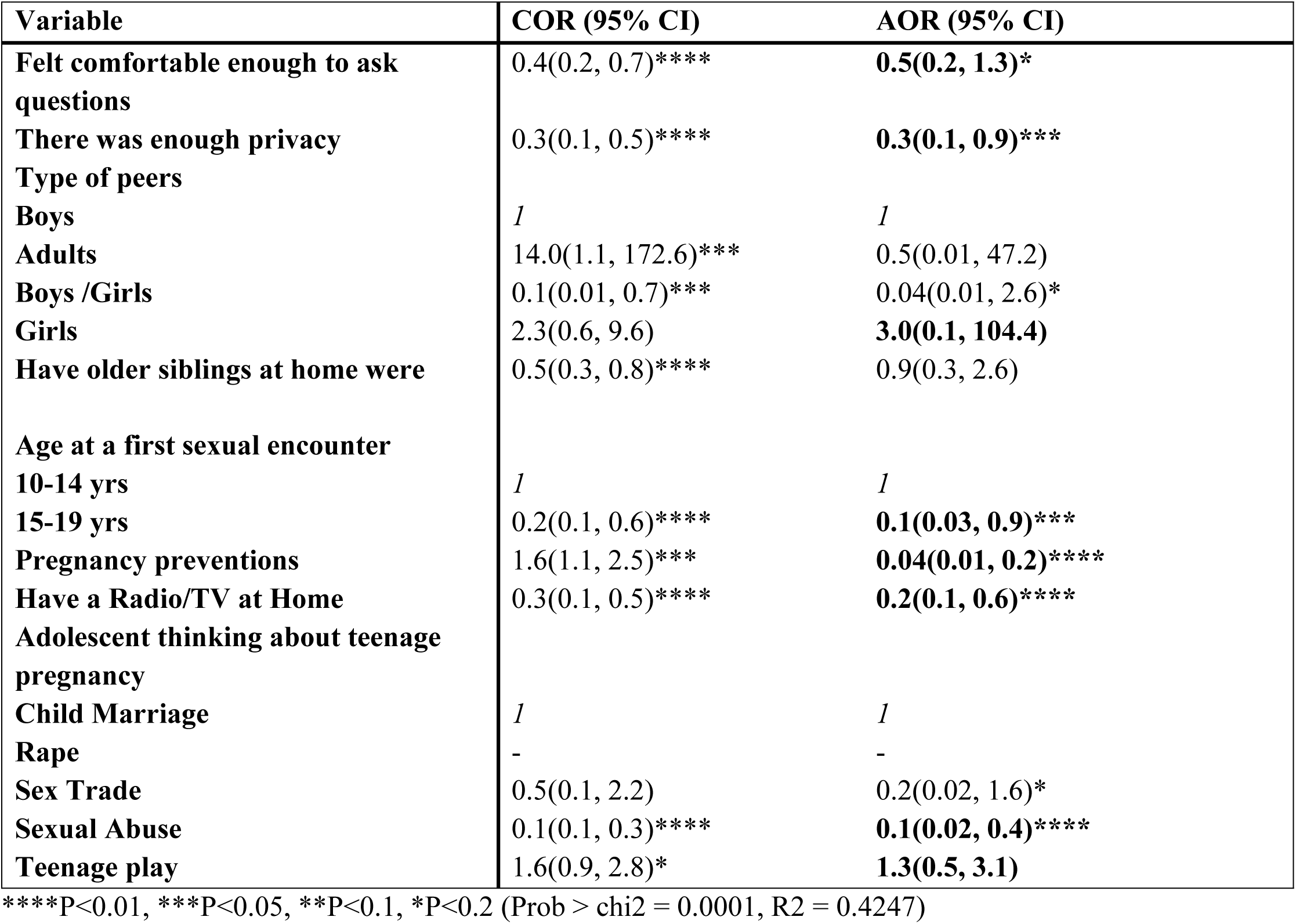
Multivariate results our adjusted variables.

## DISCUSSIONS

The findings of this study should be considered as the first examination of factors for teenage pregnancy during a COVID-19 pandemic situation in Uganda. Consequently, comprehending its implications necessitates placing it within the context of a health emergency that mandates lockdown as a public health measure for prevention. In this study, the majority (99.7%) of the girls were older teenagers (15-19 years). Even though pregnancy is often linked to an adult female, it was surprising to find many teenagers pregnant at very young ages. In this study, age was found to be statistically insignificant regarding teenage pregnancy, which might be contrary to expectations. However, initiating sexual intercourse at a later age (15–19) was associated with a reduced likelihood of becoming pregnant. This observation could be linked to the fact that teenagers in this age group may better understand their body changes and are more adept to utilizing pregnancy prevention methods. This findings agrees with a study conducted in Ethiopia(10), which also found age at first sexual intercourse to be associated teenage pregnancy. It however, contradicts what Ochen and others found in Lira, Northern Uganda (18), where age at first sex was found not to be significantly associated with teenage pregnancy.

Surprisingly, our data revealed a shift in teenage pregnancies trend from being high among secondary school attendees to high among primary school attendees. However, this was contrary to what (19), found among school-going learners in South Africa, where teenage pregnancies were higher among secondary school attendees than their primary counterparts. Although, the high level of sexual experimentation and maturity associated with secondary school teenagers supported their findings at the time. In this study, we think that the increased age at primary school graduation which is somewhere between 16 -17 years and the amount of time taken to complete primary level education in our study population could have contributed to this shift. Of all the factors investigated in this study, only having females as friends had an association with teenage pregnancies during the COVID-19 lockdown.

Conversely however, having a radio/TV at home, age at the first sexual encounter, describing teenage pregnancy as sexual abuse, feeling comfortable to ask questions during a consultation at the facility, having enough privacy during a consultation, and using pregnancy preventions were associated with less likelihood of getting pregnant.

We found that when a teenage girl associates primarily with other females as her peers, it is linked to an increased risk of teenage pregnancy during lockdown. Most of the pregnant girls in this study had friends who were exclusively female, which contradicts our understanding of female friendships. However, we believe this might be attributed to the urge to mimic what their peers are doing with boys. Nonetheless, this observation strongly hints at the potential effects of sending girls to single-sex schools as opposed to mixed-gender schools.

However, owning a radio/ TV at home during the COVID-19 lockdown was found to be associated with a reduced risk of getting pregnant. We reckon this could have been because girls who had a radio/TV at home never had to look outside their home environment for entertainment and movies during the lockdown and hence they didn’t get pregnant. This finding however agrees with another study done in South Africa among sexually active black teenagers in Cape Town (21), that found having a radio/TV at home to be associated with reduced risk of pregnancy in teenagers.

Even though there were restricted visits to the health facilities during the lockdown, we also found that, teenagers who visited health facilities during the lockdown in search for services like STIs testing /treatments, abortions, and others had reduced risk of getting pregnant.

We also found feeling comfortable to ask questions, having enough privacy, and getting a good reception from the attending clinicians while at the facility were some of the health system factors associated with less likelihood of getting pregnant.

While many pregnancy-related issues in teenagers are well-documented, including adverse obstetric outcomes, ongoing research continues to investigate various aspects of teenage pregnancy. This study, in particular, centers on birth outcomes, school dropout rates, and the location of delivery as potential factors that could predict unfavorable pregnancy outcomes in teenagers. We found that the pregnancy outcomes in these teenagers characterized by high live birth at 88.3%, with 11.1% neonatal death and 0.6% abortion in this study. Health facility delivery as a predictor of good birth outcomes for most teenage mothers was also high at 95%, compared to home deliveries. However, when we cross-tabulated to predict where the 11.1% neonatal death occurred, we found that nearly one fourth (22.2%) of these neonatal deaths happened at home compared to only 10.5% at the health facility, which was not surprising. A multitude of factors has been advanced to explain death associated with home delivery, but we think this could have been connected to delay to seek care early due to distance and other factors which are context specific. This is true for many women, as documented by (22), who found delays in deciding the place of delivery associated with home delivery and poor birth outcomes.

School dropout was also high among teenagers who became pregnant. We think this could have been because of the need to take care of the babies or fear of stigma at school associated with being pregnant. This finding agrees with another cross-sectional study done before COVID-19 in Kibuku Town Council, Kibuku District, Eastern Uganda (23)where teenage pregnancy was strongly associated with high school dropout.

### Limitation of the study

Given the COVID-19 public health emergency specific context of our findings, the results of this study must be interpreted and generalized with caution. The scope of this study did not include adverse obstetric outcomes of teenage pregnancies and other consequences like defilement, sexually transmitted diseases like HIV/AIDs, syphilis, gonorrhea, and other outcomes. Despite all this, our study has demonstrated that having only females as peers is associated with high likelihoods of getting pregnant.

In conclusion, our study was the first to determine factors associated with teenage pregnancies during a public health emergency in Uganda. This finding is vital to designing appropriate interventions in this area by NGOs and the government. It can also act as a baseline of evidence to help speed up the implementation of the newly released guidelines for preventing teenage pregnancy and management in schools 2020.

## Data Availability

The datasets used and/or analyzed during the current study are available from the corresponding author on reasonable request.

## Lists of abbreviation

ANC: Antenatal Care
NDHS: National Demographic Heath Surveys WHO-World Health Organization
COVID-19: Corona Virus Disease 2019

## Declaration

### Ethics approval and consent to participate

The ethical approval for this study was sought from the Mbale Regional Referral REC (MRRH-2021-75) and Busitema University. Informed consents were sought from the study subjects and another consent was also obtained from the parents/guardians of a child

### Consent for publication

Not Applicable

### Authors’ contributions

The primary author contributed to this study by conceptualizing the Research design and drafting the initial manuscript. All the coauthors provided expertise in critically revising the manuscript for intellectual content. The last Author supervised the overall review, provided guidance throughout the manuscript drafting, and critically reviewed and revised the manuscript for important intellectual content. All authors have read and approved the final version of the manuscript.

## Acknowledgements

We would like to acknowledge our dear research assistants for the great work they did during the data collection process. We also thank the study participants for allowing us to take part in this study and having willingly provided the information that was required to complete this study. I also like to acknowledge the contribution of the following individuals Abeso Angella, Nawanga Jascenti, Eudu James and Ouni Patrick Diox

## References

1. Darroch JE, Woog V, Bankole A. ADDING IT UP : Costs and Benefits of Meeting the Contraceptive Needs of Teenagers. New York Guttmacher Inst [Internet]. 2016 [cited 2021 Mar 1];(May):1–16. Available from: https://www.guttmacher.org/adding-it-up

2. Sedgh G, Finer LB, Bankole A, Eilers MA, Singh S. Teenage pregnancy, birth, and abortion rates across countries: Levels and recent trends. J Adolesc Heal. 2015 Feb 1;56(2):223–30.

3. UNFPA. Motherhood in Childhood: facing the challenge of teenage pregnancy (State of the World Population-2013). United Nations Population Fund New York (NY); 2013.

4. Kassa GM, Arowojolu AO, Odukogbe AA, Yalew AW. Prevalence and determinants of teenage pregnancy in Africa: a systematic review and Meta-analysis. Reprod Health. 2018;15(1):1–17.

5. Kassa GM, Arowojolu AO, Odukogbe AA, Yalew AW. Prevalence and determinants of teenage pregnancy in Africa: A systematic review and Meta-analysis 11 Medical and Health Sciences 1117 Public Health and Health Services [Internet]. Vol. 15, Reproductive Health. BioMed Central Ltd.; 2018 [cited 2021 May 2]. p. 1–17. Available from: 10.1186/s12978-018-0640-2

6. Ahinkorah BO, Kang M, Perry L, Brooks F, Hayen A. Prevalence of first teenage pregnancy and its associated factors in sub-Saharan Africa: A multi-country analysis. PLoS One [Internet]. 2021 Feb 1 [cited 2021 Jun 18];16(2 February):e0246308. Available from: 10.1371/journal.pone.0246308

7. Papri FS. Papri. Chattagram Maa-O-Shishu Hosp Med Coll J. 2016;15(1):53–6.

8. Imamura M, Tucker J, Hannaford P, Da Silva MO, Astin M, Wyness L, et al. Factors associated with teenage pregnancy in the European Union countries: A systematic review. Eur J Public Health [Internet]. 2007 Dec 1 [cited 2021 Apr 14];17(6):630–6. Available from: https://academic.oup.com/eurpub/article-lookup/doi/10.1093/eurpub/ckm014

9. Dev Raj A, Rabi B, Amudha P, Teijlingen Edwin van R, Glyn C, Raj AD, et al. Factors associated with teenage pregnancy in South Asia: A systematic review. Heal Sci J [Internet]. 2010 Jul 14 [cited 2021 Apr 14];4(1):3–14. Available from: www.hsj.gr

10. Habitu YA, Yalew A, Bisetegn TA. Prevalence and factors associated with teenage pregnancy, northeast Ethiopia, 2017: A cross-sectional study. J Pregnancy. 2018;2018.

11. Neal S, Zö Z, Matthews Z, Frost M, Fogstad H, Camacho A V, et al. The Authors Acta Obstetricia et Gynecologica Scandinavica C 2012 Nordic Federation of Societies of. Obstet Gynecol. 2012;91:1114–8.

12. WHO 2016. Global Strategy for Women ’ s, Children ’ s and Teenagers ’ Health ( 2016-2030 ). 2017;2016–7.

13. Thaithae S, Thato R. Obstetric and perinatal outcomes of teenage pregnancies in thailand. J Pediatr Adolesc Gynecol. 2011 Dec 1;24(6):342–6.

14. Trivedi SS, Pasrija S. Teenage pregnancies and their obstetric outcomes. Trop Doct [Internet]. 2007 Apr 1;37(2):85–8. Available from: 10.1177/004947550703700208

15. Uganda Bureau of Statistics. GOVERNMENT OF UGANDA Uganda Demographic and Health Survey 2016. Udhs 2016 [Internet]. 2016 [cited 2021 Feb 7];625. Available from: www.DHSprogram.com.

16. Rosenberg M, Pettifor A, Miller WC, Thirumurthy H, Emch M, Afolabi SA, et al. Relationship between school dropout and teen pregnancy among rural South African young women. Int J Epidemiol [Internet]. 2015 [cited 2021 Feb 27];44(3):928–36. Available from: https://academic.oup.com/ije/article/44/3/928/630400

17. Campbell B, Gilmore K, Kaidbey M, Laski L, Loaiza E, Martinelli-Heckadon S, et al. Motherhood in Childhood The State of World Population 2013 The editorial team is grateful for additional insights, contribu-tions and feedback from UNFPA colleagues, including [Internet]. 2013 [cited 2021 Mar 20]. 1–134 p. Available from: www.unfpa.org

18. Ochen AM, Chi PC, Lawoko S. Predictors of teenage pregnancy among girls aged 13-19 years in Uganda: A community based case-control study. BMC Pregnancy Childbirth [Internet]. 2019 Jun 24 [cited 2021 Feb 6];19(1):1–14. Available from: https://bmcpregnancychildbirth.biomedcentral.com/articles/10.1186/s12884-019-2347-y

19. S, Panday, M. MakiwaneRanchod, C, Letsoala T. Teenage Pregnancy in South Africa: With a specific focus on school-going learners [Internet]. 2009. Available from: https://repository.hsrc.ac.za/handle/20.500.11910/4711

20. World Vision 2019. The Vıolent ttruth about teenage pregnancy; What children say. World Vis Int. 2019;20(October):20–2.

21. Klitsch M. Risk factors for teenagersex. Fam Plann Perspect [Internet]. 1994;26(4):147. Available from: http://ezproxy.stir.ac.uk/login?url=http://search.ebscohost.com/login.aspx?direct=true&db=sih&AN=9410254284&site=ehost-live

22. Moindi RO, Ngari MM, Nyambati VCS, Mbakaya C. Why mothers still deliver at home: Understanding factors associated with home deliveries and cultural practices in rural coastal Kenya, a cross-section study Global health. BMC Public Health [Internet]. 2016;16(1):1–8. Available from: 10.1186/s12889-016-2780-z

23. Manzi F, Ogwang J, Akankwatsa A, Wokali OC, Obba F, Bumba A, et al. Factors Associated with Teenage Pregnancy and its Effects in Kibuku Town Council, Kibuku District, Eastern Uganda: A Cross Sectional Study. Prim Heal Care Open Access. 2018;08(02).

24. Ministry of Education and Sports. Revised Guidelines for the Prevention and Management of Teenage Pregnancy in School settings in Uganda. 2020;1–44.

25. Goonewardene IMR, Deeyagaha Waduge RPK. Adverse effects of teenage pregnancy. Ceylon Med J. 2005;50(3):116–20.

